# White matter hyperintensities classified according to intensity and spatial location reveal specific associations with cognitive performance

**DOI:** 10.1101/2020.07.10.20149575

**Authors:** Luca Melazzini, Clare E Mackay, Valentina Bordin, Sana Suri, Enikő Zsoldos, Nicola Filippini, Abda Mahmood, Vaanathi Sundaresan, Marina Codari, Eugene Duff, Archana Singh-Manoux, Mika Kivimäki, Klaus P Ebmeier, Mark Jenkinson, Francesco Sardanelli, Ludovica Griffanti

## Abstract

White matter hyperintensities (WMHs) on T_2_-weighted images are radiological signs of cerebral small vessel disease. As their total volume is variably associated with cognition, a new approach that integrates multiple radiological criteria is warranted. Location may matter, as periventricular WMHs have been shown to be associated with cognitive impairments. WMHs that appear as hypointense in T_1_-weighted images (T_1_w) may also indicate the most severe component of WMHs. We developed an automatic method that classifies WMHs into four categories (periventricular/deep and T_1_w-hypointense/nonT_1_w-hypointense) using MRI data from 684 community-dwelling older adults from the Whitehall II study. To test if location and intensity information can impact cognition, we derived two general linear models using either overall or subdivided volumes. Results showed that periventricular T_1_w-hypointense WMHs were significantly associated with poorer performance in several cognitive tests. We found no association between total WMH volume and cognition. These findings suggest that classifying WMHs according to both location and intensity in T_1_w adds value over and above total WMH volume.

**HIGHLIGHTS:**

- Heterogeneous measures of WMHs are used in research and clinical practice.
- Location and image intensity should be considered in the assessment of WMHs.
- T_1_-hypointense WMHs were found to be associated with poorer cognitive performance.
- Sub-classes of WMHs provide promising results for translation into the clinic.

## INTRODUCTION

White matter hyperintensities (WMHs) on T_2_-weighted magnetic resonance images (MRI) are radiological signs of cerebral small vessel disease (SVD) (Wardlaw et al., 2013). They are associated with a higher incidence of stroke and dementia (Debette and Markus, 2010), mood disorders, motor impairments and urinary incontinence (LADIS Study Group, 2011). Moreover, WMHs are related to cognitive impairments, particularly executive dysfunctions and poorer psychomotor speed (Bolandzadeh et al., 2012). Whilst WMHs have acquired considerable interest in the field of translational and clinical research, the assessment and the reporting of WMH volume are often inconsistent in research studies and medical practice (Frey et al., 2019).

The optimal MRI sequence to assess WMHs is fluid-attenuated inversion recovery (FLAIR). This sequence generates T_2_-weighted images where the signal from the cerebrospinal fluid is suppressed and hyperintense regions stand out on a low intensity homogeneous background (Wardlaw at al., 2013). In research, quantification of WMHs is preferred to qualitative assessment due to higher reliability, sensitivity and objectivity of the former (De Guio et al., 2016; van den Heuvel et al. 2006) and the widespread availability of segmentation software. However, the interpretation of quantitative results and comparison between studies remain difficult due to acquisition-related differences (scanner, protocol), discrepancies between processing methods (pre-processing pipelines, method/tool used to extract WMHs measurements) and variations in the definition of what should be considered a WMH (De Guio et al., 2016). Harmonisation methods that reduce or compensate for the variability due to acquisition differences and/or processing discrepancies are being developed to enable comparisons between or pooling of MRI-derived measures from different datasets (Bertani & Bordin, 2019). Notwithstanding, the lack of a clear definition on what should be segmented as a WMH and whether some WMH sub-classes are more clinically relevant than others warrant further investigation (Frey et al., 2019; Smith et al., 2019; Wardlaw et al., 2013).

Periventricular WMHs are more strongly associated with concurrent cognitive deficits than deep ones (Bolandzadeh et al., 2012). This is in line with longitudinal studies on regional baseline WMH volumes and their association with the risk of transition from intact cognition to mild cognitive impairment and dementia (De Groot et al., 2002, Kim et al., 2015, van Straaten et al., 2008). To explain this finding, the hypothesis of reduced brain reserve in periventricular regions has been put forward (De Groot et al., 2002). Despite this evidence, it is still unclear whether periventricular and deep WMHs would constitute a continuous entity or should be considered and reported separately (DeCarli et al., 2005). If the latter is true, a single method for distinguishing types of WMHs should be adopted among the many that have been proposed (Griffanti et al., 2018).

White matter hyperintensities can either be iso- or hypointense in T_1_-weighted images (Spilt et al., 2006; Wardlaw et al., 2013). To the best of our knowledge, there are no studies performed on brain tissue samples that investigate whether there is any difference in the underlying pathology between non T_1_-hypointense *versus* T_1_-hypointense WMHs. However, clinical research in multiple sclerosis has showed that demyelinating T_1_-hypointense lesions (“black holes”) represent permanent damage to the white matter and are associated with cognitive impairments (Nowaczyk et al., 2019). In WMHs of presumed vascular origin, the co-located hypointensity in T_1_-weighted images (here on referred to as T_1_-hypointense WMHs) may indicate more severe damage to the white matter than WMHs that are visible in T_2_-weighted images without any corresponding hypointensities in the T_1_-weighted images (here on referred to as *non* T_1_-hypointense WMHs). Quantitative measures of the motion of water molecules *in vivo* using diffusion tensor imaging (DTI) metrics have shown microstructural changes in the white matter in WMH areas (Wardlaw et al., 2015). Altered DTI metrics were reported in multiple sclerosis lesions that turn into permanent “black holes” (Naismith et al., 2010). Similarly, we could expect lower fractional anisotropy and higher radial diffusivity to reflect more severe axonal and myelin damage in T_1_-hypointense WMHs than in *non* T_1_-hypointense WMHs. T_1_-hypointense WMHs could thereby represent the portion of WMHs that carries the highest clinical impact.

For the reasons outlined above, we tested the hypothesis that periventricular (rather than deep) and T_1_-hypointense (rather than non T_1_-hypointense) WMHs may indicate the most severe forms of lesion in terms of impact on cognitive function. To this purpose, we classified WMHs according to both their anatomical location and intensity in T_1_-weighted images in a large cohort of community-dwelling older adults, and studied whether this classification could provide added value on the association between WMHs and cognitive function. We implemented a method for categorising WMHs according to spatial location and intensity in T_1_-weighted images that was automatic and objective and used multiple linear regression analysis to see if these WMH sub-classes show specific associations with validated scores of cognitive functions.

Given the current limitations and discrepancies in WMHs definition, our ultimate goal was to identify which sub-class(es) are specifically linked to cognitive function. This would inform future guidelines to focus the assessment on clinically-relevant radiological criteria of WMHs beyond their total extent, with a clear and objective definition of WMHs radiological appearance and location.

## METHODS

The study sample was drawn from 800 participants of the Whitehall II imaging sub-study (Filippini et al., 2014), which is part of a larger prospective occupational cohort study of British civil servants established in 1985 (Marmot and Brunner, 2005). Ethical approval was obtained from the University of Oxford Central University Research Ethics Committee, and the UCL Medical School Committee on the Ethics of Human Research. Informed written consent was obtained from all participants. Socio-demographic, health and lifestyle variables and current cognitive function were assessed at the time of the MRI.

MRI data were acquired at the Oxford Centre for Functional MRI of the Brain (FMRIB), Wellcome Centre for Integrative Neuroimaging (University of Oxford), using a 3-T, Siemens Magnetom Verio (Erlangen, Germany) scanner with a 32-channel receive head coil from April 2012 to December 2014 (N = 550 participants) and a 3-T Siemens Prisma (Erlangen, Germany) with a 64-channel receive head-neck coil from July 2015 to December 2016 (N = 250 participants) due to a scanner upgrade. Details of acquisition protocols are shown in Zsoldos et al. (2020) and Filippini et al. (2014) and are reported in Supplementary Table S1. For the purpose of this study we used high-resolution T_1_-weighted images, FLAIR images and diffusion weighted images (DWI).

All images were processed and analysed using FMRIB Software Library (FSL) v.6.0 tools (Jenkinson et al., 2012). Participants’ T_1_-weighted and FLAIR images were skull-stripped with FSL-BET (Smith et al., 2002) and bias field corrected with FSL-FAST (Zhang et al., 2001). DWI scans were pre-processed as described in (Filippini et al., 2014) and a diffusion tensor model was fit at each voxel to obtain maps of fractional anisotropy (FA), mean diffusivity (MD), axial diffusivity (AD) and radial diffusivity (RD). T_1_-weighted and FA images were then linearly registered to the corresponding FLAIR with FSL-FLIRT (Jenkinson et al., 2001). WMHs segmentation was performed with FSL-BIANCA (Griffanti et al., 2016), using intensity features (T_1_-weighted, FLAIR and FA), local average intensities (3 voxels kernel), and spatial features (MNI coordinates obtained from the transformation between FLAIR and MNI for each subject, weighting factor of 2). To avoid scanner-specific biases in the estimates, BIANCA was trained with WMH masks manually delineated in a sub-sample of individuals scanned on the Prisma (n = 24) and Verio (n = 24) scanners and an independent sample from the UK Biobank study (n = 12). The total WMH mask included voxels exceeding a probability of 0.9 of being a WMH and located within a white matter mask as described in Griffanti et al. (2016). The total WMH volume was adjusted for the total brain volume and log transformed for statistical analysis.

We separated WMHs voxels into T_1_-hypointense and non T_1_-hypointense. To achieve this, we used FSL-FAST (Zhang et al., 2001) on T1-weighted images to perform tissue type segmentation and calculate maps of partial volume estimates (PVE) for the three classes (grey matter, white matter and cerebrospinal fluid). Due to their low-intensity values, T_1_-hypointense WMHs are classified by FAST as either grey matter or cerebrospinal fluid. We therefore classified voxels as non T_1_-hypointense WMHs the voxels within the total WMH mask where the corresponding white matter PVE was greater than 0.5. We then obtained T_1_-hypointense WMHs by subtraction.

We used a cluster-based approach to separate between periventricular and deep WMHs, similar to the “continuity to ventricle” criterion described by (Griffanti et al., 2018). To do so, we created an extended ventricle mask (i.e. a ventricle mask that extended beyond the ventricular boundaries) in the Montreal Neurological Institute (MNI) space. The extended ventricle mask consisted of the probability maps -set with a very low threshold-of the lateral ventricles, thalami and fornix bilaterally. We transformed the mask to the single-subject FLAIR space via the corresponding T_1_ using linear (Jenkinson et al., 2002) and non-linear registration (Andersson et al., 2007), and classified as periventricular WMHs the clusters that overlapped with any part of the mask. Deep WMHs were then defined by subtraction.

We finally combined the two criteria and obtained four WMH masks for the following sub-classes for each participant: periventricular T_1_-hypointense WMHs; periventricular non T_1_-hypointense WMHs; deep T_1_-hypointense WMHs; deep non T_1_-hypointense WMHs. Then, each mask was used to derive the corresponding WMH volume, which was adjusted for the total brain volume and log transformed for statistical analysis.

We performed univariate multiple linear regression using the univariate general linear model function type III sum of squares on SPSS version 25.0 (IBM Corp. Armonk, NY). The following measures of participants’ cognition were selected as indices of global functioning, executive function, processing speed, and memory, in line with (Bolandzadeh et al. 2012): Montreal cognitive assessment (MoCA), trail-making test (TMT, A and B), digit span forward, backwards and sequence, digit symbol, digit coding, Boston naming-60 test (BNT), phonemic (letter) fluency (FLU-L) and semantic (category) fluency (FLU-C) tests. For details on the cognitive tests please refer to (Filippini et al., 2014). Demographic variables (age at the examination, sex, total years of education, systolic blood pressure, diastolic blood pressure) were used as covariates of no interest. For each cognitive test (dependent variable in the univariate multiple linear regression), two models were investigated, in which participants’ demographic variables were kept unchanged while WMHs were included as either WMHs total volume (corresponding to the BIANCA output) or subdivided WMH volumes (corresponding to the four sub-classes reported above).

To better investigate the meaning of T_1_-hypointensity in our sample, we performed two *post-hoc* analyses. First, we studied WMH microstructure using DTI-derived metrics, given the lack of histopathological data for this dataset. Accordingly, we compared T_1_-hypointense and non T_1_-hypointense WMHs in terms of average fractional anisotropy (FA), mean diffusivity (MD), axial diffusivity (AD) and radial diffusivity (RD) using paired *t*-tests to evaluate potential differences in the underlying microstructure. Second, since we noticed that a WMH often includes both T_1_-hypointense and non T_1_-hypointense voxels, we adopted an alternative T_1_-weighted intensity-based classification of WMHs by dividing the total WMH map into WMH clusters with and without T_1_-hypointense voxels. This sub-classification was then used to look at the prevalence of WMH clusters with a T_1_-hypointense component and to better interpret the results of the main analysis.

We also used Pearson correlations to further investigate the associations between WMH sub-classes, total WMH volume and age.

Statistical significance was set at α=0.05 (Di Leo & Sardanelli, 2020).

## RESULTS

Participants characteristics (demographics, cognitive scores and MRI measures) are reported in Table 1. One hundred and sixteen participants were excluded from our analysis due to incomplete or poor-quality images (N = 44), neurological disorders (N = 30) and inaccurate WMH segmentation masks (N = 42) to leave a total of 684 community-dwelling older adults. An example of the WMH segmentation output resulting from the method we developed is provided in Figure 1.

**Table 1.**
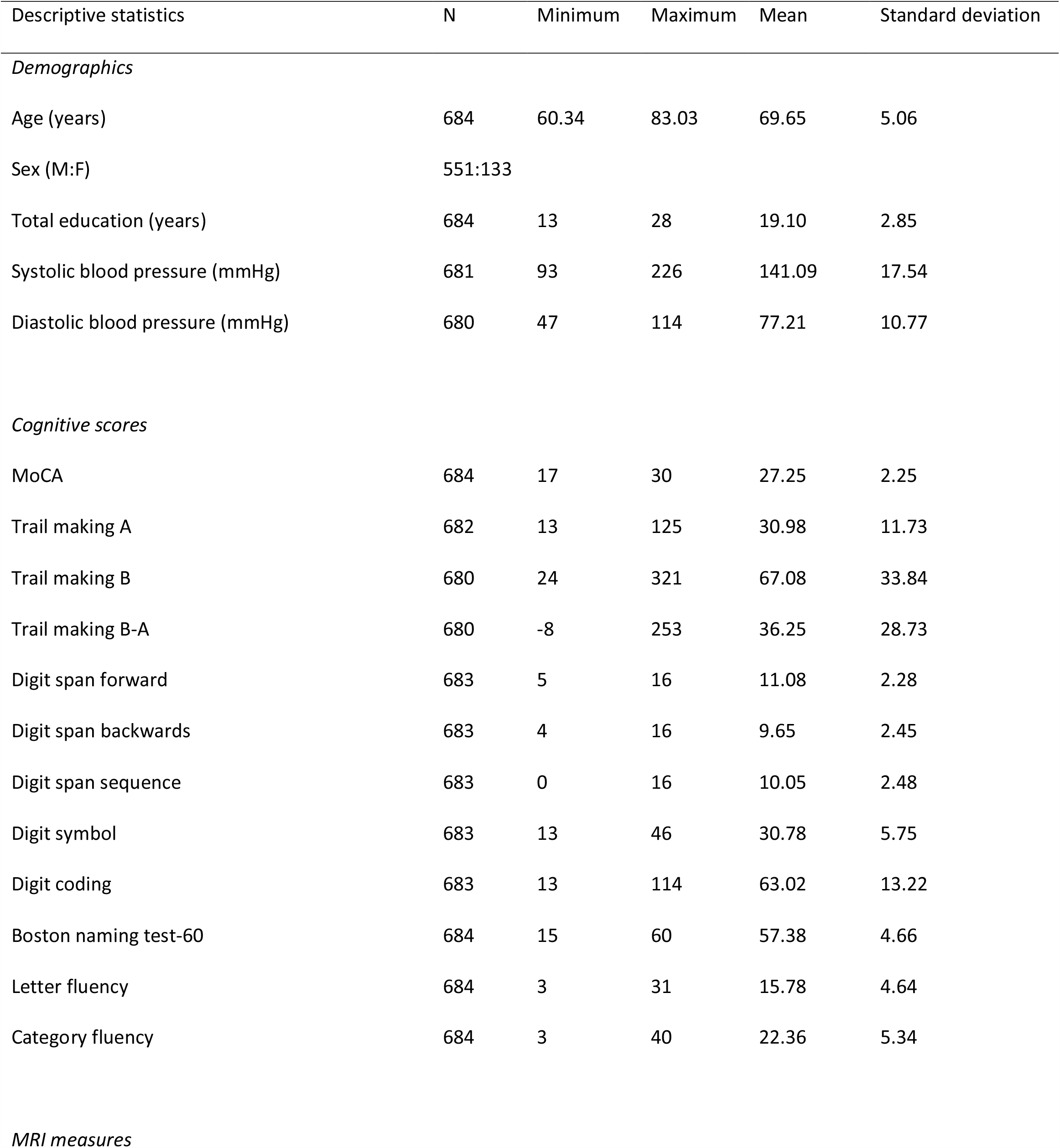
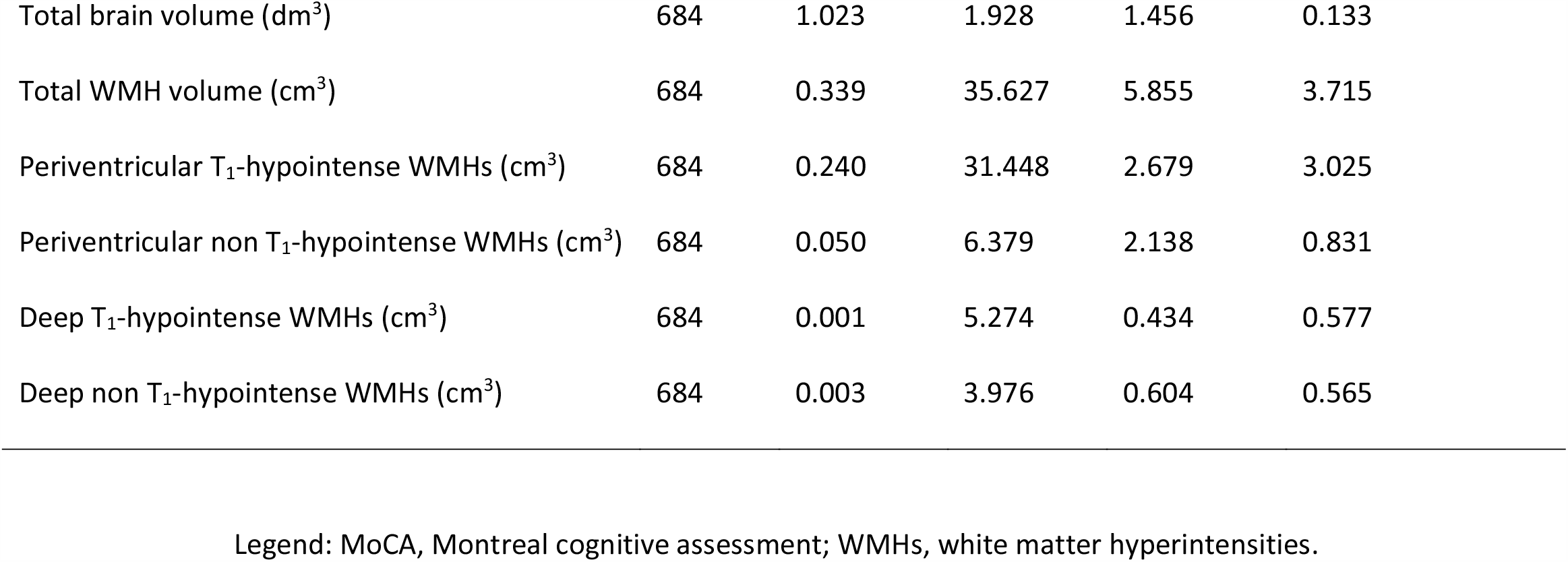
Sample characteristics.

| Descriptive statistics | N | Minimum | Maximum | Mean | Standard deviation |
| --- | --- | --- | --- | --- | --- |
| *Demographics* |  |  |  |  |  |
| Age (years) | 684 | 60.34 | 83.03 | 69.65 | 5.06 |
| Sex (M:F) | 551:133 |  |  |  |  |
| Total education (years) | 684 | 13 | 28 | 19.10 | 2.85 |
| Systolic blood pressure (mmHg) | 681 | 93 | 226 | 141.09 | 17.54 |
| Diastolic blood pressure (mmHg) | 680 | 47 | 114 | 77.21 | 10.77 |
| *Cognitive scores* |  |  |  |  |  |
| MoCA | 684 | 17 | 30 | 27.25 | 2.25 |
| Trail making A | 682 | 13 | 125 | 30.98 | 11.73 |
| Trail making B | 680 | 24 | 321 | 67.08 | 33.84 |
| Trail making B-A | 680 | -8 | 253 | 36.25 | 28.73 |
| Digit span forward | 683 | 5 | 16 | 11.08 | 2.28 |
| Digit span backwards | 683 | 4 | 16 | 9.65 | 2.45 |
| Digit span sequence | 683 | 0 | 16 | 10.05 | 2.48 |
| Digit symbol | 683 | 13 | 46 | 30.78 | 5.75 |
| Digit coding | 683 | 13 | 114 | 63.02 | 13.22 |
| Boston naming test-60 | 684 | 15 | 60 | 57.38 | 4.66 |
| Letter fluency | 684 | 3 | 31 | 15.78 | 4.64 |
| Category fluency | 684 | 3 | 40 | 22.36 | 5.34 |
| *MRI measures* |  |  |  |  |  |
| Total brain volume (dm<sup>3</sup>) | 684 | 1.023 | 1.928 | 1.456 | 0.133 |
| Total WMH volume (cm<sup>3</sup>) | 684 | 0.339 | 35.627 | 5.855 | 3.715 |
| Periventricular T<sub>1</sub>-hypointense WMHs (cm<sup>3</sup>) | 684 | 0.240 | 31.448 | 2.679 | 3.025 |
| Periventricular non T<sub>1</sub>-hypointense WMHs (cm<sup>3</sup>) | 684 | 0.050 | 6.379 | 2.138 | 0.831 |
| Deep T<sub>1</sub>-hypointense WMHs (cm<sup>3</sup>) | 684 | 0.001 | 5.274 | 0.434 | 0.577 |
| Deep non T<sub>1</sub>-hypointense WMHs (cm<sup>3</sup>) | 684 | 0.003 | 3.976 | 0.604 | 0.565 |

Legend: MoCA, Montreal cognitive assessment; WMHs, white matter hyperintensities.

**Figure 1.**
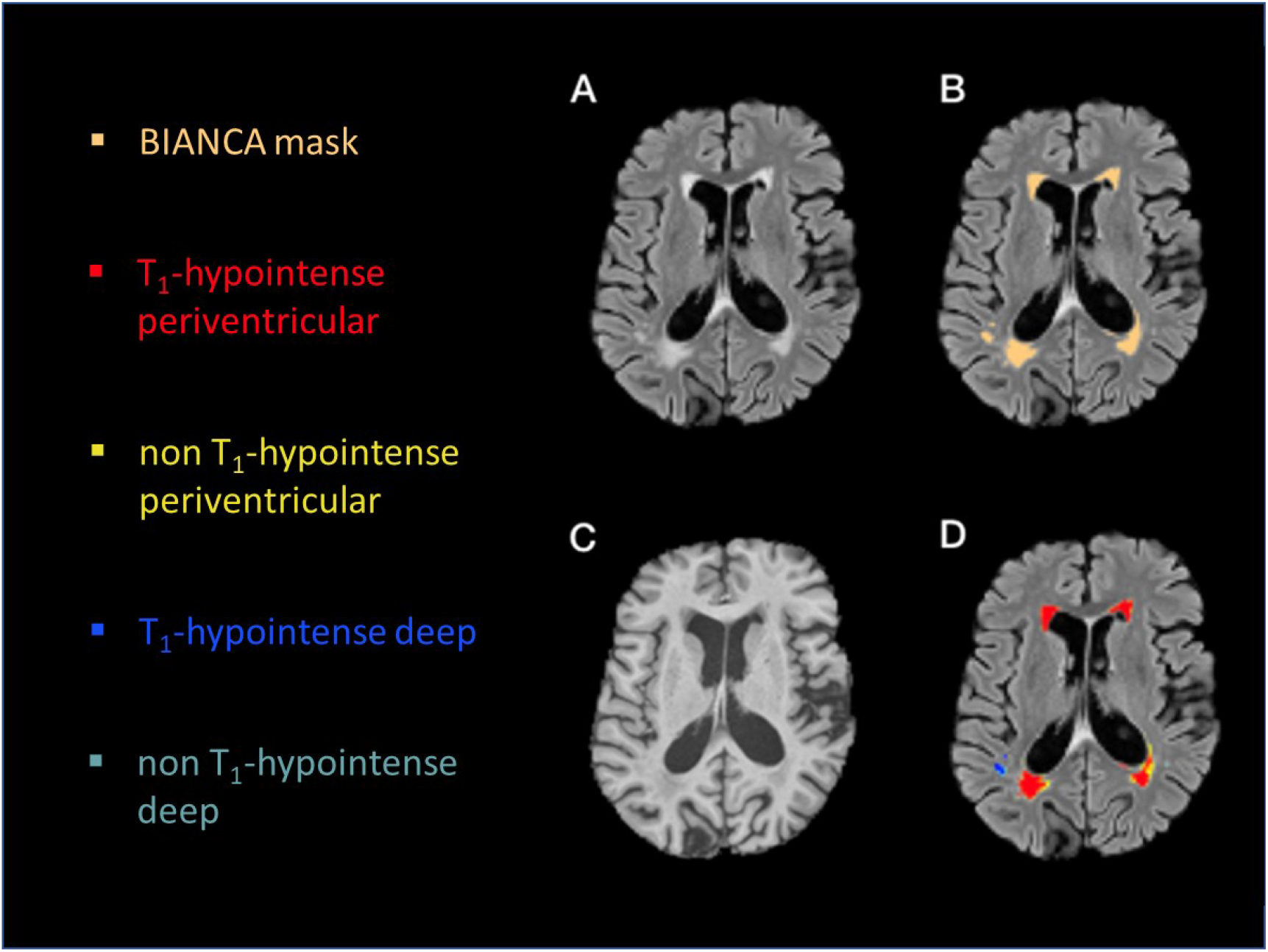
Native images and image processing output. (A) FLAIR image, (B) WMH mask (BIANCA mask - mustard colour) on FLAIR, (C) T_1_-weighted image, (D) WMH sub-classes on FLAIR: red = T_1_-hypointense periventricular WMHs; yellow = non T_1_-hypointense periventricular WMHs; blue = T_1_-hypointense deep WMHs; light blue = non T_1_-hypointense deep WMHs.

White matter hyperintensities expressed as total volume were not significantly associated with any of the cognitive tests. However, when the four sub-classes of WMHs were instead used in the model, we found statistically significant associations between periventricular T_1_-hypointense WMHs and poorer performance on the TMT-A (p = 0.011), digit symbol (p = 0.028) and digit coding (p = 0.009) tests. Conversely, non T_1_-hypointense periventricular WMHs were associated with higher scores on the digit backwards test (p = 0.023). Also, deep non T_1_-hypointense WMHs were found to be positively associated with the letter (p = 0.004) and category (p = 0.036) fluency tests. Regression analysis results for the models where WMH sub-classes were significantly associated with cognitive performances are shown in Table 2. Comprehensive results for all the models, including all covariates, are showed in Supplementary Table S2.

**Table 2.**
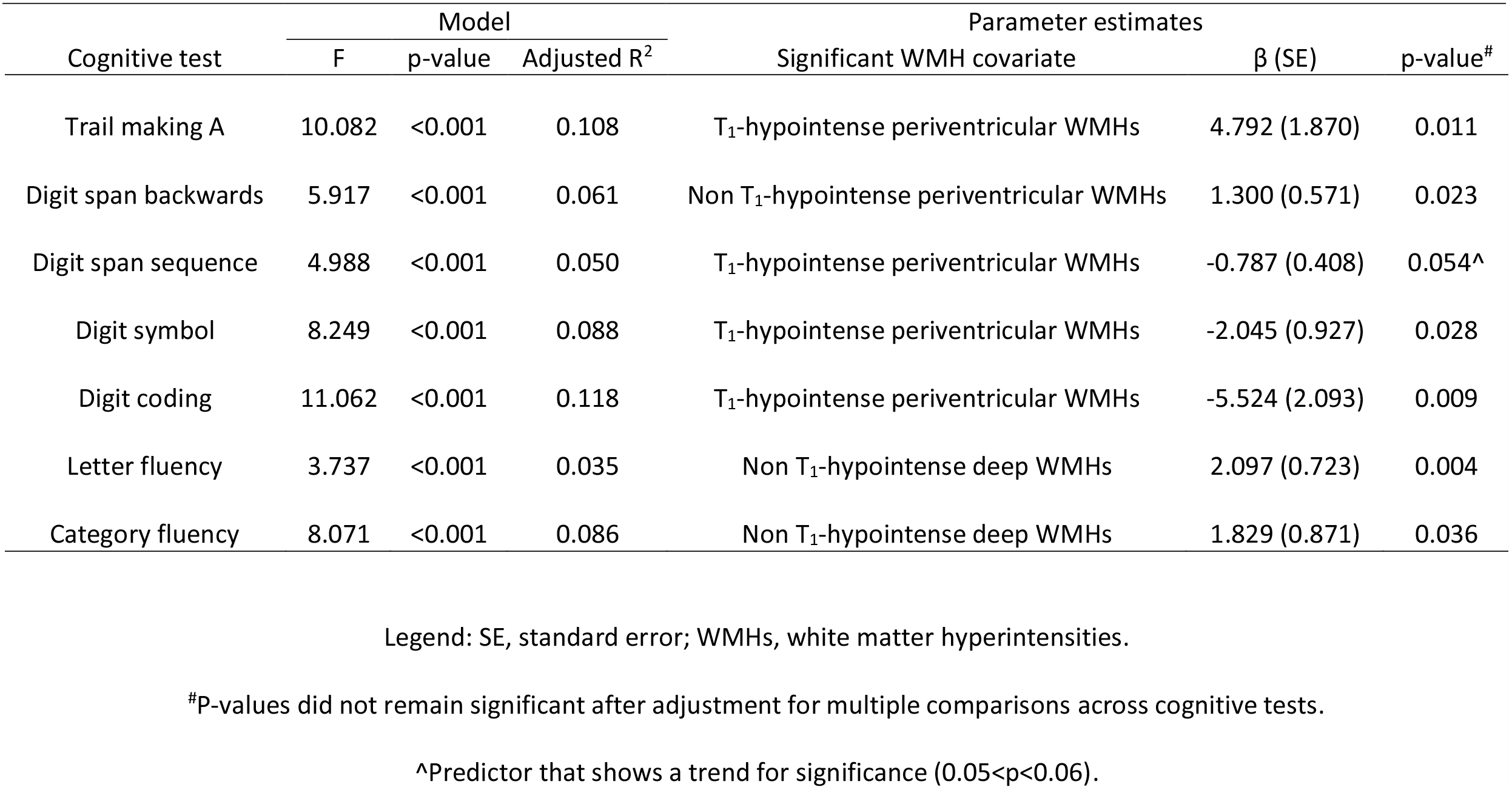
Multiple linear regression analysis results. Model results and significant WMH predictors of cognitive scores adjusted for age at the examination, sex, total years of education, systolic blood pressure and diastolic blood pressure.

| Cognitive test | F | p-value | Adjusted R<sup>2</sup> | Significant WMH covariate | β (SE) | p-value<sup>#</sup> |
| --- | --- | --- | --- | --- | --- | --- |
| Trail making A | 10.082 | <0.001 | 0.108 | T<sub>1</sub>-hypointense periventricular WMHs | 4.792 (1.870) | 0.011 |
| Digit span backwards | 5.917 | <0.001 | 0.061 | Non T<sub>1</sub>-hypointense periventricular WMHs | 1.300 (0.571) | 0.023 |
| Digit span sequence | 4.988 | <0.001 | 0.050 | T<sub>1</sub>-hypointense periventricular WMHs | -0.787 (0.408) | 0.054<sup>^</sup> |
| Digit symbol | 8.249 | <0.001 | 0.088 | T<sub>1</sub>-hypointense periventricular WMHs | -2.045 (0.927) | 0.028 |
| Digit coding | 11.062 | <0.001 | 0.118 | T<sub>1</sub>-hypointense periventricular WMHs | -5.524 (2.093) | 0.009 |
| Letter fluency | 3.737 | <0.001 | 0.035 | Non T<sub>1</sub>-hypointense deep WMHs | 2.097 (0.723) | 0.004 |
| Category fluency | 8.071 | <0.001 | 0.086 | Non T<sub>1</sub>-hypointense deep WMHs | 1.829 (0.871) | 0.036 |

Legend: SE, standard error; WMHs, white matter hyperintensities.
<sup>#</sup>P-values did not remain significant after adjustment for multiple comparisons across cognitive tests.
<sup>^</sup>Predictor that shows a trend for significance (0.05<p<0.06).

When looking at microstructural differences in WMHs classified according to the intensity in T_1_-weighted images criterion, T_1_-hypointense WMHs showed significantly lower FA, and higher MD, AD and RD than non T_1_-hypointense WMHs (p < 0.001 for all measures). Results of the within-subject paired *t*-tests for the averages of DTI-derived metrics between T_1_- and non T_1_-hypointense WMHs are reported in Table 3.

**Table 3.**
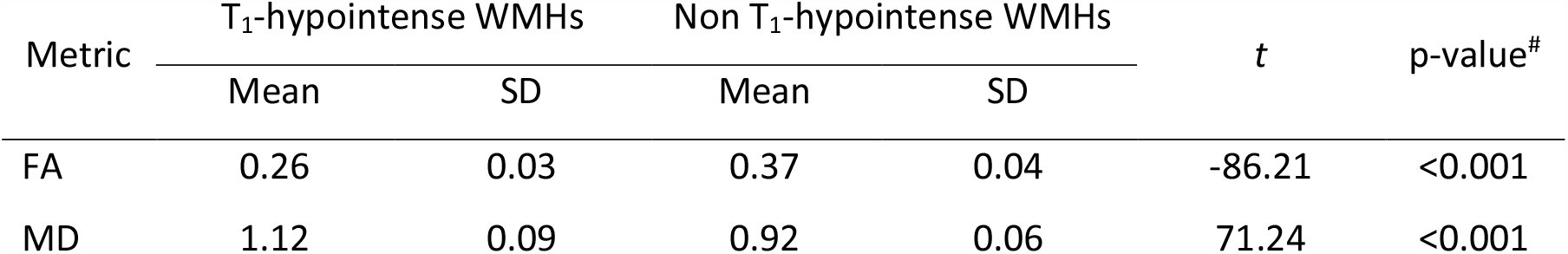
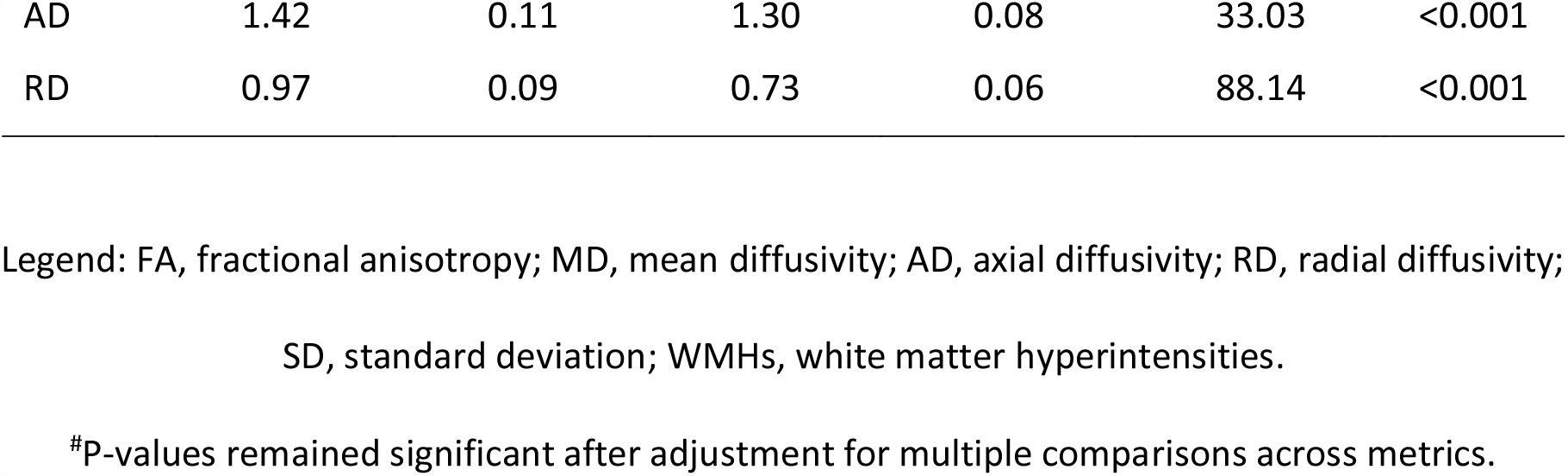
Comparison between the T_1_- and non T_1_-hypointense WMHs in terms of DTI-derived metrics. Paired *t*-tests results. Diffusivity values are expressed in (mm^2^/s) × 10^−3^.

Results for the separation of the total WMH volume into WMH clusters with and without corresponding T_1_-hypointense voxels are reported in Table 4.

**Table 4.** Number and size of WMH clusters with and without T_1_-hypointense voxels.

|  | Number | Average volume<br>*per subject* (mm<sup>3</sup>) | Average volume<br>*per cluster* (mm<sup>3</sup>) |
| --- | --- | --- | --- |
| WMH clusters | 223.91 ± 103.77 | 5855.01 ± 3715 | 35.50 ± 41.70 |
| Clusters without T<sub>1</sub>-hypointense voxels | 112.43 ± 59.43 | 295.72 ± 188.87 | 2.58 ± 0.72 |
| Clusters with T<sub>1</sub>-hypointense voxels | 111.48 ± 57.55 | 5559.29 ± 3754.63 | 66.16 ± 63.13 |
| *Rims* |  | 2446.69 ± 1021.26 | 27.96 ± 18.99 |
| *Cores* |  | 3112.61 ± 3227.86 | 49.72 ± 18.99 |

We found that 50% of the WMH clusters had T_1_-hypointense voxels. However, despite constituting half of all clusters, these WMHs composed 94% of the whole WMH volume. The remaining 6 % of the WMH volume consisted of small non T_1_-hypointense WMH clusters, i.e. WMH clusters without co-located hypointensity in T_1_-weighted images.

The volume of non T_1_-hypointense WMHs is the sum of non T_1_-hypointense clusters and the hyperintense rims of T_1_-hypointense clusters (a schematic representation is depicted in Figure 2).

**Figure 2.**
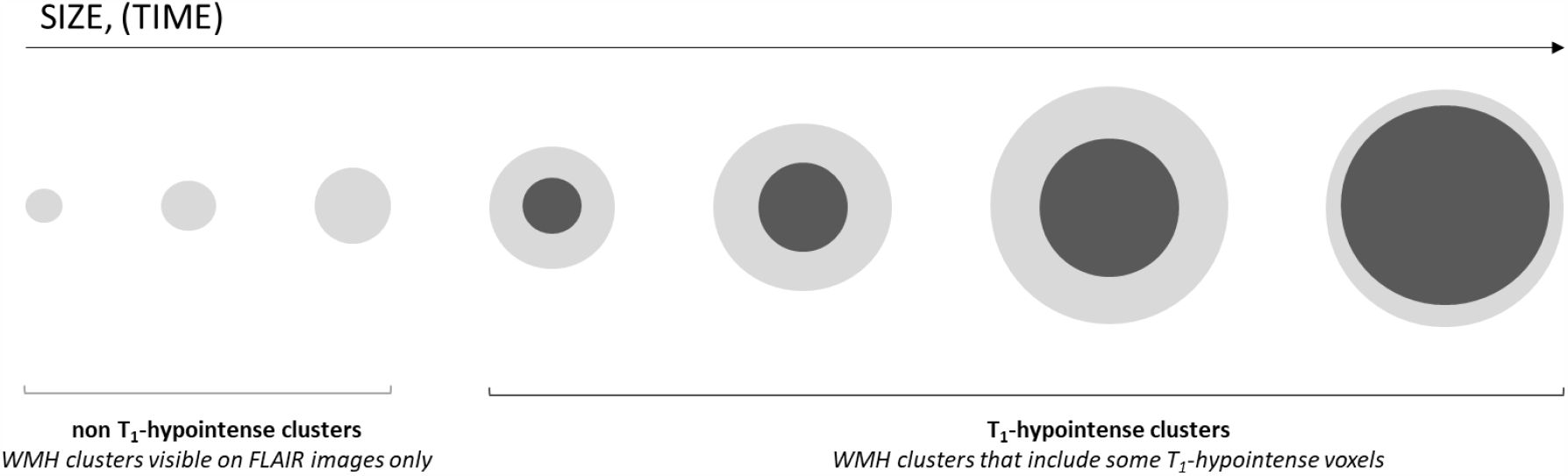
Diagram representing the potential evolution of WMHs over time. WMHs, white matter hyperintensity.

Given the unexpected significant positive associations of non T_1_-hypointense WMHs with the digit span backwards, letter and category fluency, we investigated which of these two components (either non T_1_-hypointense clusters or rims of T_1_-hypointense clusters) drove the observed associations. When fitting multiple linear regression models with either of these two components as the only WMH predictor for the fluency and digit span backwards scores, we found that non T_1_-hypointense clusters were significantly associated with the fluency performances (FLU-L: β = 1.976, p = 0.001; FLU-C: β = 2.023, p = 0.004). Conversely, rims were significant predictors of the digit span backwards scores (β = 0.916, p = 0.035).

Additional multiple regression models and Pearson correlations among WMH sub-classes, total WMH volume and age supporting these findings are reported in Supplementary Table S3 and S4.

## DISCUSSION

In this study we sought to provide a clinically-oriented insight into WMHs by developing an automated method for classifying WMHs according to spatial location (periventricular *versus* deep WMHs) and lesion intensity in the corresponding T_1_-weighted image (T_1_-hypointense *versus* non T_1_-hypointense WMHs). We fitted univariate multiple linear regression models using the volumes of WMH sub-classes as predictors for the participants’ performance in several cognitive tests and then further explored the microstructural properties of T_1_-hypointense WMHs to understand the meaning of this radiological appearance. Our classification proved to be clinically meaningful, as periventricular T_1_-hypointense WMHs were found to be linked to poorer performance on multiple cognitive tests, including the trail making A, digit symbol and digit coding tests. Our results suggest that periventricular T_1_-hypointense WMHs would represent the best WMH biomarker for cognitive impairment.

Our finding of periventricular WMHs being associated with poorer cognition is in line with previous large longitudinal population-based studies on non-demented elderly (de Groot et al., 2002; Griffanti et al., 2018), where periventricular WMHs were found to be more strongly associated with participants’ cognition than deep WMHs. Increased volume of periventricular WMHs in the Alzheimer Disease Neuroimaging Initiative dataset was also associated with evidence of beta-amyloid deposition in the brain (Marnane et al., 2016), suggesting a synergistic damage driven by concurrent SVD and Alzheimer’s pathology in these areas. Periventricular regions are also characterised by high density of long associating fibres which link the cortex to the deep grey matter and other distant brain territories (Filley 1998). For this reason, they are potentially susceptible to pathologies that damage cortical arteries and eventually provoke distal hypoperfusion (Moody et al., 1990). Disrupted cholinergic activity is related to periventricular (and not deep) WMHs and may be involved in the physio-pathological pathway that underpins the observed cognitive scores (Bohnen et al., 2009). Moreover, high periventricular WMHs were found to be associated with frontal cortical thinning, where both imaging findings were independently linked to executive dysfunctions (Seo et al., 2012). Altogether, these findings endorse our results of periventricular WMHs being more strongly associated with cognitive impairment than deep WMHs.

Although the location criterion is well established, less is known about the meaning of T_1_-intensity in WMHs. This aspect has been well-investigated in demyelinating disease where hypointense lesions in T_1_-weighted images are more likely to represent low axonal density and irreversible tissue damage (Bitsch et al., 2001; van Walderveen et al., 1998). Our DTI analysis showed decreased FA and increased MD, AD and RD in WMHs T_1_-hypointense WMHs. Not surprisingly, these findings mirror similar microstructural results found in multiple sclerosis (Vavasour et al.,2018) and suggest more severe damage to the white matter in T_1_-hypointense areas than their T_1_-isointense counterpart.

T_1_-hypointense and non T_1_-hypointense WMHs may represent two distinct entities with different meanings. Alternatively, WMHs could start as small punctate FLAIR hyperintensities and later develop a T_1_-hypointense “core”. Despite the lack of longitudinal data in our cohort, we attempted to investigate further the meaning of the intensity in T_1_-weighted images and the theoretical evolution of WMHs and their intensity in T_1_-weighted images from a cross-sectional basis. Within the total WMH mask, we separated WMH clusters that contained some T_1_-hypointense voxels (T_1_-hypointense clusters) from those that did not (non T_1_-hypointense clusters) (Table 4). Since most of the WMH volume comprised (relatively big) clusters with a T_1_-hypointense component, our results suggest that the evolution of a WMH may be a cascade of events starting from small punctate lesions leading to bigger lesions with a T_1_-hypointense core and surrounding non T_1_-hypointense rim (Figure 2).

This mechanism could explain the significant association we observed between higher volume of deep non T_1_-hypointense WMHs and higher scores at the fluency tests. Since the volume of non T_1_-hypointense WMHs includes both the volume of non T_1_-hypointense clusters and the outer rim of the clusters with a T_1_-hypointense core, we hypothesized that the positive association of deep non T_1_-hypointense WMHs with higher cognitive scores at the fluency tests could be driven by non-T_1_ hypointense clusters (i.e. smaller, less severe WMHs, that have not yet progressed into a WMH with a T_1_-hypointense component). We tested this hypothesis by fitting multiple linear regression models with either the volume of all non T_1_-hypointense clusters or only the volume of deep non T_1_-hypointense WMHs voxels as the only imaging predictors for the fluency tests (Supplementary Table S3). Notably, non T_1_-hypointense clusters were more strongly associated with fluency scores than deep non T_1_-hypointense WMHs. Thus, it is likely that the volume of clusters of non T_1_-hypointense WMHs drove the observed significance of deep non T_1_-hypointense WMHs as predictors of better cognitive scores at the fluency tests.

The positive associations of non T_1_-hypointense clusters with fluency scores could be due to the fact that these small lesions are more frequent in younger individuals with a low WMH volume. The hypothesized evolution into T_1_-hypointense clusters would then explain the apparent decrease of this type of lesions in individuals with lower fluency scores.

Since non T_1_-hypointense clusters are small, we cannot exclude the possibility that some of these are false positives from BIANCA segmentation. However, WMH masks have been visually checked so that we exclude results that could be driven by errors in WMHs segmentation.

Our finding of higher non T_1_-hypointense periventricular WMHs linked to higher scores at the digit span backwards test could instead be explained by the other component of non T_1_-hypointense WMHs, the non T_1_-hypointense voxels belonging to T_1_-hypointense clusters (i.e. the hyperintense “rims”). In fact, when fitting the multiple linear regression model with the volumes of rims of all T_1_-hypointense clusters as the only imaging predictor, we found a positive association with the digit span backwards that was very similar to the one given by non T_1_-hypointense periventricular WMHs alone (Supplementary Table S3). Although rim and core belong to the same physical entity (i.e. the WMH cluster) they are likely to have opposite meanings. On the one hand, T_1_-hypointense WMHs voxels predict bad cognitive scores, as seen for the Trail Making Test A, digit symbol and digit coding tests. On the other hand, in spite of the positive association between rims and cores in terms of overall volume, the former are predictors of higher cognitive scores from the digit span backwards test. Thus, within the context of our hypothesized evolution of WMHs, rims would represent those WMH areas belonging to T_1_-hypointense clusters that have not turned T_1_-hypointense yet and theoretically “withstand” further tissue damage. When this occurs, the number of non T_1_-hypointense voxels would decrease because they become T_1_-hypointense. This would in turn explain the positive relationship with cognition observed in our results. Further investigation would be necessary to explore the potential cascade of events leading to change in T_1_ intensity in a longitudinal setting, since the hypothesis of rims as WMHs associated with healthy cognitive aging is very speculative.

Our study has some limitations in terms of the data and the methodology.

The Whitehall II imaging sub-study dataset shows a strong gender imbalance, skewed towards men, since it reflects the demographic of British civil servants at the time of recruitment in the main study. Moreover, the age range of the sample is quite narrow (60-84 years). Future work is therefore needed to test the generalisability of our results. Our approach for the sub-classification of WMHs relies on automated segmentation of WMHs with BIANCA, tissue type segmentation with FAST, and registration of images with different spatial resolutions. As already mentioned, we cannot exclude inaccuracies in the WMH masks, despite visual inspection of the results. We used FAST segmentation as a proxy for defining T_1_-hypointensity, therefore inaccuracies in the segmentation would translate to inaccuracies in the sub-classification. Moreover, we performed linear and non-linear registrations between images with different resolutions (FLAIR, T_1_ and MNI space) and the interpolation process could have slightly affected the segmented volumes.

Finally, our hypothesis on the evolution of WMHs should be interpreted cautiously and prompt further longitudinal studies. For example, it would be very valuable to follow up participants and study how WMH sub-classes evolve over time to validate the proposed theory. Another interesting future development would be looking at how the different WMH sub-classes are related to incidence of disease using risk models in well-balance longitudinal datasets.

Despite these limitations, this study presents some novel theoretical and methodological insights that can contribute to better understanding of the role of WMHs in cognitive aging. The methods developed herein can be easily adopted in other research settings. The extended ventricle mask and the scripts created for images post-processing are publicly available^1^. These scripts can also be equally applied to any manually- or automatically-derived WMH masks, other than those from BIANCA, to obtain the four WMH sub-classes presented in this study.

In conclusion, we showed that information from spatial location and intensity in T_1_-weighted images provides potentially clinically useful insights into the meaning of WMHs with regards to participants’ cognitive function. Notably, the combination of these two criteria revealed an association with cognitive scores related to global functioning, executive function, processing speed, and memory that the WMH total volume alone could not provide.

## Data Availability

The study follows MRC data sharing policies (https://www.mrc.ac.uk/research/policies-and-guidance-for-researchers/data-sharing/). Data will be accessible via the Dementias Platform UK (https://portal.dementiasplatform.uk/) after 2020. The extended ventricle mask described in the methods section and the scripts created for images post-processing are already publicly available (https://git.fmrib.ox.ac.uk/ludovica/wmh-sub-classes).

## ACKNOWLEDGMENTS

We thank all Whitehall II participants for their time, the Whitehall II staff at the University College London, Mandy Pipkin and Barbora Krausova for assisting with recruitment and data collection, the FMRIB Radiographers team for data acquisition, IT and support teams at the Wellcome Centre for Integrative Neuroimaging for their helpful collaboration. The study follows MRC data sharing policies (https://www.mrc.ac.uk/research/policies-and-guidance-for-researchers/data-sharing/). Data will be accessible via the Dementias Platform UK (https://portal.dementiasplatform.uk/) after 2020.

## Footnotes

1 Git repository: https://git.fmrib.ox.ac.uk/ludovica/wmh-sub-classes

